## Supplementary tables 2-6 - ROB tables for "The role of SARS-CoV-2 variants of concern in children and adolescents with COVID-19: a systematic review"

| Supplementary table 2: Risk of bias assessment - Child/adolescent symptoms/severity of disease when infected with a VOC | | | | | | | | |
| --- | --- | --- | --- | --- | --- | --- | --- | --- |
| **Cohort studies^[1]^** | **Exposed and non-exposed cohorts from the same population** | **Confidence in exposure assessment** | **Confidence that outcome of interest was not present at start of study** | **Matching of exposed and unexposed for all variables associated with outcome of interest or adjustment of prognostic variables by statistical analysis** | **Confidence in assessment of presence/absence of prognostic factors** | **Confidence in outcome assessment** | **Adequate follow-up of cohorts** | **Similar co-interventions between groups** |
| Ulyte et al. [15] | definitely yes | probably no | definitely no | probably no | probably yes | probably no | definitely no | definitely yes |
| Nakel et al. [18] | probably yes | definitely yes | probably yes | definitely no | definitely no | probably no | definitely no | probably yes |
| Somekh et al. [19] | probably yes | definitely yes | probably yes | probably no | probably no | definitely yes | probably yes | probably no |
| Swann et al. [20] | definitely yes | definitely yes | probably no | probably yes | definitely yes | definitely yes | probably yes | probably yes |
| Edward et al. [35] | probably yes | definitely yes | definitely yes | probably yes | probably no | definitely yes | probably yes | probably no |
| Celebi et al. [34] | probably yes | probably yes | definitely yes | definitely no | definitely no | probably yes | probably no | definitely no |
| Stewart et al. [22] | probably yes | definitely yes | probably yes | probably no | probably no | definitely yes | definitely yes | probably no |
| Brookman et al. [21] | probably yes | definitely yes | definitely yes | definitely no | definitely no | probably yes | probably no | probably yes |
| Li et al. [23] | definitely yes | definitely yes | probably yes | definitely yes | probably no | probably yes | definitely yes | definitely yes |
| Shoji et al. [29] | definitely yes | definitely yes | definitely yes | probably no | probably yes | definitely yes | probably yes | probably no |
| Fisman et al. [28] | probably yes | definitely yes | definitely yes | probably yes | probably yes | definitely yes | probably yes | probably yes |
| Ryu et al. [27] | probably yes | definitely yes | probably yes | definitely no | definitely no | probably yes | probably no | probably no |
| Martin et al. [30] | probably yes | probably yes | definitely yes | probably no | probably no | definitely yes | probably no | definitely no |
| Molteni et al. [32] | definitely yes | probably yes | probably yes | probably yes | probably no | probably yes | probably yes | probably no |
| Murillo-Zamora et al. [26] | probably yes | definitely yes | definitely yes | definitely no | probably no | definitely yes | probably no | probably no |
| Butt et al. [31] | probably yes | probably yes | probably yes | probably yes | probably no | definitely yes | probably no | probably no |
| Oliveira et al. [47] | probably yes | definitely yes | definitely yes | definitely no | definitely no | definitely yes | probably no | probably no |
| Cloete et al. [36] | probably yes | probably yes | probably yes | probably yes | probably yes | probably yes | definitely yes | definitely yes |
| Wang et al. [41] | probably yes | probably yes | probably yes | probably yes | probably yes | definitely yes | probably no | probably no |
| Wang et al. [42] | probably yes | probably yes | probably yes | probably no | probably no | definitely yes | probably no | probably no |
| Marks et al. [43] | definitely yes | definitely yes | definitely yes | definitely no | definitely no | probably no | definitely yes | probably no |
| Butt et al. [39] | probably yes | definitely yes | probably yes | probably yes | probably yes | definitely yes | definitely no | probably yes |
| Kildegaard et al. [33] | probably yes | probably yes | probably yes | definitely no | probably no | definitely yes | definitely yes | probably yes |
| Fowlkes et al. [40] | definitely yes | probably yes | probably yes | probably no | definitely no | probably yes | probably yes | probably yes |
| Hao et al. [24] | probably yes | probably yes | probably yes | probably no | definitely no | probably yes | probably yes | probably yes |
| Brewster et al. [37] | definitely yes | definitely yes | definitely yes | definitely no | definitely yes | definitely yes | probably yes | probably no |
| Waltenburg et al. [17] | probably yes | probably yes | probably yes | probably no | probably no | probably yes | definitely no | probably yes |
| Marks et al. [46] | probably yes | probably no | probably yes | probably no | probably no | definitely yes | probably yes | probably yes |
| Nygaard et al. [25] | probably yes | probably yes | definitely yes | probably no | definitely no | definitely yes | definitely yes | probably yes |
| **Case control studies^[2]^** | **Confidence in exposure assessment** | **Confidence in cases but not controls acquiring outcome of interest** | **Proper selection of cases  (those that were exposed and developed outcome of interest)** | **Proper selection of controls  (those that were exposed and did not develop outcome of interest)** | **Matching of cases and controls according to  prognostic variables or statistical adjustment for these variables** |  |  |  |
| Roberts et al. [14] | probably yes | probably yes | definitely yes | probably yes | definitely no |  |  |  |
| **Cross sectional studies** | **Exposed and non-exposed groups from the same population** | **Confidence in exposure assessment** | **Confidence that outcome of interest was not present at start of study** | **Matching of exposed and unexposed for all variables associated with outcome of interest or adjustment of prognostic variables by statistical analysis** | **Confidence in assessment of presence/absence of prognostic factors** | **Confidence in outcome assessment** | **Similar co-interventions between groups** |  |
| Iuliano et al. [44] | probably yes | probably yes | definitely yes | definitely no | definitely no | definitely yes | probably no |  |
| Shi et al. [45] | definitely yes | definitely yes | probably yes | definitely no | definitely yes | definitely yes | probably yes |  |
| Somekh et al. [16] | probably yes | definitely yes | probably no | definitely no | probably no | definitely yes | probably yes |  |

| Supplementary table 3: Risk of bias assessment - Child/Adolescent risk of severe disease when infected with a VOC | | | | | | | | |
| --- | --- | --- | --- | --- | --- | --- | --- | --- |
| **Cohort studies^[3]^** | **Exposed and non-exposed cohorts from the same population** | **Confidence in exposure assessment** | **Confidence that outcome of interest was not present at start of study** | **Matching of exposed and unexposed for all variables associated with outcome of interest or adjustment of prognostic variables by statistical analysis** | **Confidence in assessment of presence/absence of prognostic factors** | **Confidence in outcome assessment** | **Adequate follow-up of cohorts** | **Similar co-interventions between groups** |
| Butt et al. [39] | probably yes | definitely yes | probably yes | probably yes | probably yes | definitely yes | definitely no | probably yes |
| **Cross sectional studies** | **Exposed and non-exposed groups from the same population** | **Confidence in exposure assessment** | **Confidence that outcome of interest was not present at start of study** | **Matching of exposed and unexposed for all variables associated with outcome of interest or adjustment of prognostic variables by statistical analysis** | **Confidence in assessment of presence/absence of prognostic factors** | **Confidence in outcome assessment** | **Similar co-interventions between groups** |  |
| Shi et al. [45] | definitely yes | definitely yes | probably yes | definitely no | definitely yes | definitely yes | probably yes |  |

| Supplementary table 4: Risk of bias assessment - Child/Adolescent risk of becoming infected with a VOC | | | | | | | | |
| --- | --- | --- | --- | --- | --- | --- | --- | --- |
| **Cohort studies^[4]^** | **Exposed and non-exposed cohorts from the same population** | **Confidence in exposure assessment** | **Confidence that outcome of interest was not present at start of study** | **Matching of exposed and unexposed for all variables associated with outcome of interest or adjustment of prognostic variables by statistical analysis** | **Confidence in assessment of presence/absence of prognostic factors** | **Confidence in outcome assessment** | **Adequate follow-up of cohorts** | **Similar co-interventions between groups** |
| Nakel et al. [18] | probably yes | definitely yes | probably yes | definitely no | definitely no | probably no | definitely no | probably yes |
| Somekh et al. [19] | probably yes | definitely yes | probably yes | probably no | probably no | definitely yes | probably yes | probably no |
| Abu Raddad et al. [54] | definitely yes | definitely yes | probably no | definitely no | probably no | probably yes | definitely yes | probably no |
| Loenenbach et al. [52] | probably yes | probably yes | probably no | definitely no | definitely no | definitely yes | probably no | probably no |
| Lorthe et al. [53] | definitely yes | definitely yes | probably yes | definitely no | probably yes | probably yes | definitely yes | definitely yes |
| Clifford et al. [55] | probably yes | probably yes | definitely yes | definitely no | definitely no | definitely yes | probably no | probably yes |
| Dougherty et al. [57] | probably yes | definitely yes | definitely yes | probably no | probably yes | definitely yes | probably yes | probably no |
| Singanayagam et al. [58] | probably yes | probably yes | probably yes | probably no | probably no | definitely yes | probably yes | probably no |
| Schenk et al. [50] | probably yes | probably yes | probably no | probably no | probably no | probably yes | probably no | probably yes |
| Lorthe et al. [59] | probably yes | definitely yes | definitely no | definitely no | definitely no | definitely yes | probably no | definitely no |
| Waltenburg et al. [17] | probably yes | probably yes | probably yes | probably no | probably no | probably yes | definitely no | probably yes |
| Ng et al. [56] | probably yes | probably yes | probably yes | probably no | definitely no | probably yes | probably yes | probably yes |
| **Case control studies^[5]2^** | **Confidence in exposure assessment** | **Confidence in cases but not controls acquiring outcome of interest** | **Proper selection of cases  (those that were exposed and developed outcome of interest)** | **Proper selection of controls  (those that were exposed and did not develop outcome of interest)** | **Matching of cases and controls according to  prognostic variables or statistical adjustment for these variables** |  |  |  |
| Roberts et al. [14] | probably yes | probably yes | definitely yes | probably yes | definitely no |  |  |  |
| **Cross sectional studies** | **Exposed and non-exposed groups from the same population** | **Confidence in exposure assessment** | **Confidence that outcome of interest was not present at start of study** | **Matching of exposed and unexposed for all variables associated with outcome of interest or adjustment of prognostic variables by statistical analysis** | **Confidence in assessment of presence/absence of prognostic factors** | **Confidence in outcome assessment** | **Similar co-interventions between groups** |  |
| Somekh et al. [16] | probably yes | definitely yes | probably no | definitely no | probably no | definitely yes | probably yes |  |
| Neuberger et al. [51] | probably yes | probably yes | probably yes | probably no | probably no | probably yes | probably yes |  |

| Supplementary table 5: Risk of bias assessment - Child/adolescent risk of transmission when infected with a VOC | | | | | | | | |
| --- | --- | --- | --- | --- | --- | --- | --- | --- |
| **Cohort studies^[6]^** | **Exposed and non-exposed cohorts from the same population** | **Confidence in exposure assessment** | **Confidence that outcome of interest was not present at start of study** | **Matching of exposed and unexposed for all variables associated with outcome of interest or adjustment of prognostic variables by statistical analysis** | **Confidence in assessment of presence/absence of prognostic factors** | **Confidence in outcome assessment** | **Adequate follow-up of cohorts** | **Similar co-interventions between groups** |
| Somekh et al. [19] | probably yes | definitely yes | probably yes | probably no | probably no | definitely yes | probably yes | probably no |
| Buchan et al. [65] | definitely yes | probably yes | probably no | definitely no | definitely no | probably yes | probably no | probably no |
| Lindstrom et al. [60] | definitely yes | definitely yes | definitely no | probably no | definitely no | probably yes | probably no | probably yes |
| Loenenbach et al. [52] | probably yes | probably yes | probably no | definitely no | definitely no | definitely yes | probably no | probably no |
| Lorthe et al. [53] | definitely yes | definitely yes | probably yes | definitely no | probably yes | probably yes | definitely yes | definitely yes |
| Julin et al. [63] | probably yes | definitely yes | definitely yes | probably no | definitely no | definitely yes | definitely yes | probably yes |
| Lyngse et al. [61] | definitely yes | definitely yes | definitely yes | probably yes | probably yes | probably yes | probably yes | probably no |
| Chudasama et al. [62] | probably yes | probably no | probably no | definitely no | definitely no | probably yes | probably no | definitely no |
| Lorthe et al. [59] | probably yes | definitely yes | definitely no | definitely no | definitely no | definitely yes | probably no | definitely no |
| Trobajo-Sanmartin  et al. [68\| | probably yes | definitely yes | probably yes | probably yes | probably yes | definitely yes | probably no | probably yes |
| Waltenburg et al. [17] | probably yes | probably yes | probably yes | probably no | probably no | probably yes | definitely no | probably yes |
| Ng et al. [56] | probably yes | probably yes | probably yes | probably no | definitely no | probably yes | probably yes | probably yes |

| **Case control studies^[7]2^** | **Confidence in exposure assessment** | **Confidence in cases but not controls acquiring outcome of interest** | **Proper selection of cases  (those that were exposed and developed outcome of interest)** | **Proper selection of controls  (those that were exposed and did not develop outcome of interest)** | **Matching of cases and controls according to  prognostic variables or statistical adjustment for these variables** |
| --- | --- | --- | --- | --- | --- |
| Loss et al. [64] | probably yes | definitely yes | definitely yes | definitely yes | probably no |
| Allen et al. [67] | definitely yes | definitely yes | definitely yes | definitely yes | probably yes |

| **Modelling studies^[8]3^** | **Is the population relevant?** | **Are any critical interven-tions missing?** | **Are any relevant out-comes missing?** | **Is the context (settings and circum-stances) applicable?** | **Is external validation of the model sufficient to make its results credible for your decision?** | **Is internal verification of the model sufficient to make its results credible for your decision?** | **Does the model have sufficient face validity to make its results credible for your decision?** | **Is the design of the model adequate for your decision problem?** | **Are the data used in  populating the model suitable for your decision problem?** | **Were the analyses performed using the model adequate to inform your decision problem?** | **Was there an adequate assessment of the effects of uncertainty?** | **Was the reporting of the model adequate to inform your decision problem?** | **Was the interpre-tation of results fair and balanced?** | **Were there any  potential conflicts of interest?** | **If there were potential  conflicts of interest, were steps taken to address these?** |
| --- | --- | --- | --- | --- | --- | --- | --- | --- | --- | --- | --- | --- | --- | --- | --- |
| Ratman et al. [66] | definitely yes | probably yes | probably no | probably yes | definitely no | definitely no | probably no | probably yes | probably yes | probably yes | definitely no | probably yes | probably yes | definitely no | probably no |

| Supplementary table 6: Risk of bias assessment - Risk of long-term effects when infected with a VOC | | | | | | | | |
| --- | --- | --- | --- | --- | --- | --- | --- | --- |
| **Cohort studies^[9]7^** | **Exposed and non-exposed cohorts from the same population** | **Confidence in exposure assessment** | **Confidence that outcome of interest was not present at start of study** | **Matching of exposed and unexposed for all variables associated with outcome of interest or adjustment of prognostic variables by statistical analysis** | **Confidence in assessment of presence/absence of prognostic factors** | **Confidence in outcome assessment** | **Adequate follow-up of cohorts** | **Similar co-interventions between groups** |
| Molteni et al. [28] | definitely yes | probably yes | probably yes | probably yes | probably no | probably yes | probably yes | probably no |
| Kildegaard et al. [xx] | probably yes | probably yes | probably yes | definitely no | probably no | definitely yes | definitely yes | probably yes |
