## Supplementary tables 7-12 - tables of studies for "The role of SARS-CoV-2 variants of concern in children and adolescents with COVID-19: a systematic review"

| **Supplementary table 7: Review question 1 - Child/adolescent symptoms/severity of disease when infected with the alpha variant (n=9)** | | | | | | | | | | |
| --- | --- | --- | --- | --- | --- | --- | --- | --- | --- | --- |
| **Author** | **Study design** | **Country** | **Study setting** | **Number of study participants <20 yo^[1]^** | **Number of adult study participants** | **Age groups** | **Mean/ Median age** | **VOC^[2]^** | **Symptoms/Severity** | **Comments** |
| Roberts  et al. [14] | case-control study | United Kingdom | schools kindergartens crèches | overall: 83  infected: 21 | 34 | ≤4yo, adults | n/r | Alpha | **Symptoms**  24% fever, 19% cough (52% asymptomatic) | **sequencing:** 1 case was sequenced, rest was assumed based on date of sampling and national prevalence |
| Ulyte  et al.  [15] | cohort study | Switzerland | schools kindergartens crèches | overall:  2 487  newly  infected: 182 | n/a | 7-17 yo | median:  12 yo | Alpha | **Symptoms**  17% fatigue (30/182), 17% sore throat (30/182), 15% headache (27/182), 14% fever (26/182) | **sequencing:** n/r |
| Somekh  et al. [16] | cross-sectional study | Israel | households | alpha  infection: 31  non-VOC infection: 20 | n/a | 11-17 yo | n/r | Alpha | **Symptoms (alpha vs. non-VOC)** sensory impairment: 6.5% (2/31) vs. 40% (8/20) (RR^[3]^: 0.16; 95%CI^[4]^: 0.04–0.6) | **sequencing:** n/r |

| Waltenburg et al.  [17] | cohort study | USA^[5]^ | households | n/r | n/r | <5 yo, 5-11 yo, 12-17 yo | median:  12 yo  (IQR^[6]^ 7-15) | Alpha | **Symptoms (alpha vs. non-VOC)**  constitutional (chills, fever, myalgia): OR^[7]^ 5.5 (95%CI: 1.55-19.20)  lower respiratory (e.g. cough): OR 3.52 (95%CI: 1.26-9.86)  gastrointestinal (e.g. diarrhoea, pain): OR 3.55 (95%CI: 1.16-10.89)  **Severity (alpha vs. non-VOC):**  Symptom duration (median, days):  3 vs. 2, p^[8]^ = 0.64 | **vaccination status:** 97% unvaccinated (147/151), 3% partially vaccinated (4/151)  **sequencing:** majority of primary cases sequenced |
| --- | --- | --- | --- | --- | --- | --- | --- | --- | --- | --- |
| Nakel  et al. [18] | cohort study | Germany | schools kindergartens crèches | infected: 40  alpha  infection: 18  non-VOC infection: 22 | n/a | ≤6 yo | n/r | Alpha | **Symptoms (alpha vs. non-VOC)** fever: 38.9% (7/18) vs. 50% (11/22) cough: 44.4% (8/18) vs. 36.4% (8/22) shortness of breath: 11.1% (2/18) vs. 0% (0/22)  **Severity (alpha vs. non-VOC)** ICU^[9]^ admission: 0 vs. 0 | **sequencing:** partially |
| Somekh  et al. [19] | cohort study | Israel | hospital inpatients  hospital outpatients | alpha infection:  72 796  non-VOC infection:  26 689 | n/a | ≤9 yo | n/r | Alpha | **Severity (alpha vs. non-VOC)** Hospitalisation: 0.52% (379/72796) vs. 0.98% (261/26689) RR 0.53 (95% CI: 0.46-0.63), p < 0.001  Severe disease/death: 6.9% (26/379) vs. 6.5% (17/261) RR 0.99 (95% CI: 0.96-1.04) | **sequencing:** n/r |
| Swann  et al. [20] | cohort study | United Kingdom | hospital inpatients | infected: 1 617  alpha  infection: 952  non-VOC infection: 665 | n/a | ≤19 yo | median (alpha):  6.5 yo  (IQR 0.3-14.9)  median (non-VOC): 4.0 yo  (IQR 0.4-13.6) | Alpha | **Symptoms (alpha vs. non-VOC)** fever: 57.1% (544/952) vs. 73.8% (491/665), p<0.001  **Severity (alpha vs. non-VOC)** Length of hospitalisation: 2 days (IQR 1-4) vs. 2 days (IQR 1-4), p=0.079 critical care: 12.0% (109/910) vs. 12.7 (78/616), p=0.989 | **sequencing:** n/r |
| Brookman et al.  [21] | cohort study | United Kingdom | hospital outpatients | infected: 80  alpha  infection: 60  non-VOC infection: 20 | n/a | ≤18 yo | n/r | Alpha | **Severity (alpha vs. non-VOC)** Critical disease: 3% (2/60) vs. 20% (4/20) Oxygen: 8% (5/60) vs. 35% (7/20) non-invasive ventilation: 3% (2/60) vs. 15% (3/20) invasive ventilation: 2% (1/60) vs. 20% (4/20) | **sequencing:** n/r |
| Stewart  et al. [22] | cohort study | United Kingdom | hospital inpatients | infected: 110  alpha  infection: 67 non-VOC infection: 43 | n/a | <18 yo | median overall:  10.2 yo  (IQR 7.6-12.6) | Alpha | **Symptoms (alpha vs. non-VOC)** PIMS-TS^[10]^: 67 vs. 43 cases acute kidney injury in PIMS-TS:  28.4% (19/67) vs. 33% (14/43), p=0.85 | **sequencing:** n/r |

| **Supplementary table 8: Review question 1 - Child/adolescent symptoms/severity of disease when infected with the delta variant (n=13)** | | | | | | | | | | |
| --- | --- | --- | --- | --- | --- | --- | --- | --- | --- | --- |
| **Author** | **Study design** | **Country** | **Study setting** | **Number of study participants <20 yo^[11]^** | **Number of adult study partici-pants** | **Age groups** | **Mean/ Median age** | **VOC^[12]^** | **Symptoms/Severity** | **Comments** |
| Li  et al. [23] | cohort  study | China | hospital inpatients | infected: 77 | n/a | <12 yo | median: 9 yo | Delta | **Symptoms**  75.3% fever (58/77), 18.2% cough (14/77), 7.8% expectoration (6/77) | **sequencing:** all everyone was hospitalised for isolation |
| Hao  et al.  [24] | cohort  study | China | hospital inpatients | infected: 66 | n/a | ≤18 yo | mean (±SD^[13]^):  10.35±5.86 yo | Delta | **Symptoms**  47.0% fever (31/66), 47.0% cough (31/66), 34.8% nasal congestion (23/66), 25.8% sore throat (17/66), 10.6% fatigue (7/66), 4.5% decreased sense of smell (3/66) | **Sequencing:** n/r |
| Nygaard  et al.  [25] | cohort  study | Denmark | hospital inpatients | infected: 74  delta infection : 51  non-VOC infection: 23 | n/a | ≤17 yo | n/r | Delta | **MIS-C**^[14]^ **symptoms (delta vs. non-VOC)** hypotension/shock: 51% (26/51) vs. 57% (13/23) cardiac involvement: 90% (46/51) vs. 100% (23/23) gastrointestinal involvement: 98% (50/51) vs. 100% (23/23) respiratory involvement: 12% (6/51) vs. 26% (6/23) **MIS-C severity (delta vs. non-VOC)** ICU^[15]^ admission: 55% (28/51) vs. 52% (12/23) vasoactive support: 16% (8/51) vs. 22% (5/23) no need for mechanical ventilation or ECMO^[16]^, no deaths for both delta and non-VOC infection  (risk of MIS-C comparable with delta and non-VOC infection) | **vaccination status:** none vaccinated **sequencing:** majority sequenced |
| Murillo-Zamora  et al.  [26] | cohort  study | Mexico | hospital inpatients | infected:  26 961  person-days:  297 099 | n/r | ≤19 yo | mean (±SD): 14.4±5.8 yo | Delta | **Severity (delta vs. pre-delta)** pneumonia: incidence rate 30.3/10 000 person-days vs. 9.4/10 000 person-days RR^[17]^ 0.98 (95%CI: 0.97-0.99), p^[18]^<0.001 | **sequencing:** n/r no information on pre-delta strains |
| Ryu  et al.  [27] | cohort  study | South Korea | population | infected: 235  delta infection:  92  pre-delta infection:  143 | n/r | <18 yo | mean (delta):  10.2 yo  mean  (pre-delta):  13.8 yo p<0.001 | Delta | **Symptoms (delta vs. pre-delta)** rhinorrhoea: 10.5% (15/143) vs. 25% (23/92), p=0.003 nasal stuffiness: 15.4% (22/143) vs. 34.8% (32/92), p=0.001 sore throat: 12.6% (18/143) vs. 23.9% (22/143), p=0.02  no difference in prevalence of other COVID symptoms (e.g. fever, cough, pneumonia) | **sequencing:** n/r no information on pre-delta strains |
| Fisman  et al.  [28] | cohort  study | Canada | population | infected: 20 434  delta infection: 12 387  non-VOC infection: 8 047 | n/a | ≤19 yo | n/r | Delta | **Severity (delta vs. non-VOC)** hospitalization (<10 yo): OR^[19]^ 2.51 (95%CI: 1.23-5.11) hospitalization (10-19 yo): OR 0.78 (95%CI: 0.38-1.91)   ICU admission (10-19 yo): OR 1.37 (95%CI: 0.11-16.46) | **sequencing:** all sequenced |
| Shoji  et al. [29] | cohort  study | Japan | hospital inpatients | infected: 1 299  delta infection: 349  pre-delta infection: 950 | n/a | <18 yo | median (delta):  7.0 yo  (IQR^[20]^: 2-13)  median (pre-delta):  10.0 yo  (IQR: 4-15) p<0.001 | Delta | **Severity (delta vs. pre-delta)** ICU admission: 1.4% (5/349) vs. 0.1% (1/950), p=0.006 length of hospitalisation (median, days): 7.0 (IQR 5.0-9.0) vs. 8.0 (IQR 5.0-9.0), p=0.031 mechanical ventilation: 0% vs. 0% | **sequencing:** n/r no information on pre-delta strains |
| Martin  et al.  [30] | cohort  study | USA^[21]^ | hospital inpatients hospital outpatients | infected: 167 262  delta infection: 29 191  pre-delta infection: 138 071 | n/a | <19 yo | median (delta):  10.3 yo  (IQR 5.3-14.9)  median (pre-delta):  12.3 yo  (IQR 6.2-16.4) | Delta | **Severity (delta vs. pre-delta)** hospitalisation: 6.0% (1738/29191) vs. 6.2% (8507/138071), p=0.18 severe disease: 10.3% (179/1738) vs. 14.6% (1242/8507), (OR 0.67, 95%CI: 0.57-0.79), p<0.001 severe disease (children classified as non-black, non-white): OR 1.69 (95%CI: 1.02-2.82, p=0.04 | **sequencing:** n/r no information on pre-delta strains |
| Butt  et al. [31] | cohort  study | Qatar | population | infected: 246  delta infection: 125  beta infection: 121 | n/a | ≤19 yo | n/a | Beta, Delta | **Symptoms (delta vs. beta)** moderate disease: aOR^[22]^ 0.17 (0.06-0.48) vs. 0.35 (0.09-1.39), p=0.54 severe disease: aOR 0.08 (0.01-0.48) vs. NA, p>0.99 | **sequencing:** all sequenced |
| Molteni  et al. [32] | cohort  study | United Kingdom | population | infected: 1 400  delta infection: 706 5-11 yo: 227 12-17 yo:  479  alpha infection: 694 5-11 yo:  276 12-17 yo:  418 | n/r | 5-17 yo | median (alpha):  12 yo  (IQR 9-15)  median (delta):  14 yo  (IQR 13-16) | Alpha, Delta | **Symptoms (delta vs. alpha)** 5-11yo  headache: 60.8% (138/227) vs. 39.9% (110/276), OR 2.31 (95%CI: 1.6-3.33), p<0.001 fever: 46.3% (105/227) vs. 31.9% (88/276), OR 1.89 (95%CI: 1.31-2.73) (p=0.001) 12-17yo headache: 73.7% (353/479) vs. 61.5% (257/418), OR 1.81 (95%CI: 1.36-2.42), p<0.001 fever: 43.2% (207/479) vs. 27.0% (113/418), OR 2.05 (95%CI: 1.55-2.72), p<0.001 rhinorrhoea: 57.6% (276/479) vs. 47.4% (198/418), OR 1.52 (95%CI: 1.17-1.98), p=0.002 sore throat: 60.5% (290/479) vs. 45.7% (191/418), OR 1.87 (95%CI: 1.43-2.45), p<0.001  **Severity (delta vs. alpha)** symptom burden (5-11 yo, median/days): 4 (IQR 2-7) vs 3 (IQR 2-5) symptom burden (12-17 yo, median/days): 6 (IQR 3-9) vs 5 (IQR 3-8) | **sequencing:** n/r |
| Kildegaard et al.  [33] | cohort  study | Denmark | Hospital inpatients and outpatients | infected: 74 611  alpha infection: n/r  delta infection: n/r | n/a | <18 yo | Median (infected):  8 yo  (IQR 1-14) | Alpha, Delta | **Severity (alpha vs. delta)**  hospitalization: RR^[23]^ 0.49% (98/20102) (95%CI^[24]^: 0.4-0.59) vs. 0.48% (18/16694) (95%CI: 0.38-0.6)  MIS-C: RR 0.04% (9/20164) (95%CI: 0.02-0.08) vs. 0.04% (5/12778) (95%CI: 0.01-0.09) | **vaccination status:** partially vaccinated  **sequencing:** n/r |
| Çelebi  et al. [34] | cohort  study | Turkey | hospital inpatients hospital outpatients | infected: 680  alpha infection: 329  beta/gamma infection: 17  delta infection: 165  non-VOC infection: 169 | n/a | ≤17 yo | median: 9.25 yo | Alpha, Beta, Delta | **Severity  (alpha vs. delta vs. beta/gamma vs. non-VOC)** hospitalization: 9.4% (31/329) vs. 19.4% (32/165) vs. 18% (3/17) vs. 10.1% (17/169)   **Severity (alpha vs. delta)** hospitalization: 9.4% (31/329) vs. 19.4% (32/165), p=0.016  **Severity (delta vs. non-VOC)**  hospitalization: 19.4% (32/165) vs. 10.1% (17/169), p=0.002 | **sequencing:** all sequenced, but no differentiation between gamma and beta |
| Edward  et al.  [35] | cohort  study | USA | hospital outpatients | infected: 499    alpha infection: 96   gamma infection: 38   delta infection: 119   non-VOC infection: 243 | n/a | ≤18 yo | median (alpha):   9 yo  (IQR 2-14)   median (gamma):  7 yo  (IQR 1-14)   median (delta): 4 yo  (IQR 1-11)   median (non-VOC):  7 yo (IQR 1-12) | Alpha, Delta, Gamma | **Symptoms  (alpha vs. gamma vs. delta vs. non-VOC)**  symptomatic: 91.6% (88/96) vs. 89.4% (34/38) vs. 89.9% (107/119) vs. 91.7% (223/243), p=0.92  **Severity - Hospitalization**  non-VOC: 3.2% (8/243) reference   alpha: 7.2% (7/96): OR 1.9 (95%CI: 0.59-6.3)  gamma: 15.7% (6/38): OR 5.9  (95%: 1.6-21.5), p=0.007  delta: 6.7% (8/119): OR 2.2 (95%CI: 0.69-7.7)     **ICU admission**  non-VOC: 1.2% (3/243): reference  alpha: 1.0% (1/96) :OR 0.47  (95%CI: 0.02-4.0)  gamma: 5.2% (2/38): OR 2.6  (95%CI: 0.31-17.5)  delta: 2.5% (3/119): OR 1.4 (95%CI: 0.24-9.0)    **Respiratory support**  non-VOC: 1.2% (3/243): reference  alpha: 0% : OR 0  gamma: 10.5% (4/38) : OR 8.3  (95%CI: 1.5-56.3), p=0.02  delta: 2.5% (3/119) : OR 2.2  (95%CI: 0.34-16.9) | **sequencing:** all sequenced |

| **Supplementary table 9: Review question 1 - Child/adolescent symptoms/severity of disease when infected with the omicron variant (n=11)** | | | | | | | | | | |
| --- | --- | --- | --- | --- | --- | --- | --- | --- | --- | --- |
| **Author** | **Study design** | **Country** | **Study setting** | **Number of study participants <20 yo^[25]^** | **Number of adult study partici-pants** | **Age groups** | **Mean/ Median age** | **VOC^[26]^** | **Symptoms/severity** | **Comments** |
| Cloete  et al. [36] | cohort study | South Africa | hospital inpatients | infected: 139 | n/a | ≤13 yo | n/r | Omicron | **Symptoms** 47% fever, 40% cough, 24% vomiting, 23% difficulty breathing, 20% convulsions  **Severity** hospitalization: 92% standard ward care, 25% oxygen therapy, 2% high flow oxygen,  6% ventilation | **sequencing:** partially sequenced |
| Brewster  et al.  [37] | cohort study | USA^[27]^ | hospital inpatients, hospital outpatients | infected: 75  omicron:  61  pre-omicron: 14 | n/a | n/r | median (omicron): ED^[28]^: 2.4 yo (0.8–2.6)  hospitalized: 1.6 yo  (0.7–1.1)  median (pre-omicron): ED: 2.6 yo  (1.6–3.4) hospitalized: 1.3 yo  (1.1–1.5) | Omicron | **Symptoms (omicron vs. pre-omicron):** Croup: 81% (61/75) vs. 19% (14/75)   Median weekly croup cases (omicron vs. pre-omicron): 11 [IQR^[29]^ 2–17] vs. 0 (IQR 0-0), p^[30]^ < 0.001  none required invasive ventilation or died (irrespective of variant) | **sequencing:** n/r |
| Ludvigsson et al. [38] | case series  (>1 case) | Sweden | hospital inpatients | infected: 3 | n/r | <15 yo | n/r | Omicron | **Symptoms Case 1:** repeated convulsions over several hours, hospitalisation: 4 days **Case 2:** status epilepticus, hospitalisation: 2 days **Case 3:** one convulsion (30-60sec), short behavioural change, hospitalisation: 2 days | **sequencing:** n/r 14yo: clinical diagnosis |
| Butt  et al.  [39] | cohort study | Qatar | population | Infected: 985   (both the omicron and delta cohort) | n/r | <18 yo | median (delta):  7 yo  (IQR 3-9)  median (omicron):  6 yo  (IQR 3-10) | Delta, Omicron | **Severity (omicron vs. delta)**  mild/not hospitalised: 97.8% (963/985) vs. 84.2% (829/985), p<0.001 moderate disease: 2.2% (22/985) vs. 15.7% (155/985) critical disease: 0% (0/985) vs. 0.1% (1/985)    moderate+critical disease: 2.2% (22.985) vs. 15.8% (156/985), p<0.001,  aOR^[31]^ 0.12 (95%CI: 0.07-0.18) | **vaccination status:** none vaccinated  **sequencing:** n/r |
| Fowlkes  et al.  [40] | cohort study | USA | population | infected: 252  omicron infection: 150  delta infection: 102 | n/r | 5-11 yo, 12-15 yo | n/r | Delta, Omicron | **Symptoms (omicron vs. delta)** COVID^[32]^- symptoms: 48.7% (73/150) vs. 65.7% (67/102); OR^[33]^ 2.0 (95%CI^[34]^: 1.2-3.45), p=0.008**‘  Severity (omicron vs. delta)** total days of symptoms: 5.4% vs. 8.0%; OR -3.4 (95%CI: (-)5.7 - (-)1.0), p=0.006 hours of missed school: 21.8% vs. 24.1%**;** OR -10.6 (95%CI: (-)18.6 - (-)2.7), p=0.01  (no differences in febrile symptoms, received medical care, days spent sick in bed) | **vaccination status:**  5-11 yo: 65% (682/1052) double vaccinated, 7% (69/1052) 1 dose, and 29% (301/1052) unvaccinated.  12–15 yo: 68% (212/312) double vaccinated, 5% (15/312) 1 dose, 27% (85/312) unvaccinated **sequencing:** n/r |
| Wang  et al. [41] | cohort study | USA | hospital inpatients hospital outpatients | infected: 7 198   (both the omicron and the delta cohort) | n/r | <5 yo | mean (omicron): 1.49±1.42   mean (delta): 1.48±1.42 | Delta, Omicron | **Severity (omicron vs. delta)** ED visits: 18.83 %(1355/7198) vs. 26.67 (1920/7198); RR^[35]^ 0.71 (95% CI: 0.66-0.75) hospitalisation: 1.04% (75/7198) vs. 3.14 (226/7198); RR 0.33 (95%CI: 0.26-0.43) ICU^[36]^ admission: 0.14% (10/7198) vs. 0.43% (31/7198); RR 0.32 (95%CI: 0.16-0.66) mech. ventilation: 0.33% (24/7198) vs. 1.15% (83/7198); RR 0.29 (95%CI: 0.18-0.46) | **sequencing:** n/r |
| Wang  et al. [42] | cohort study | USA | hospital inpatients hospital outpatients | infected ≤4 yo: 11 556 (both the omicron and the delta cohort)  infected  5-17 yo: 18 715 (both the omicron and the delta cohort) | n/a | ≤4 yo, 5-17yo | n/a | Delta, Omicron | **Severity (omicron vs. delta)** ≤ 4yo ED visits: 20.41% (2359/11556) vs. 23.96% (2796/1156); RR 0.85 (95%CI: 0.81-0.89) hospitalization: 1.55% (179/11556) vs. 3.0% (347/11556); RR 0.52 (95%CI: 0.43-0.62) ICU admissions: 0.1% (12/11556) vs. 0.42% (49/11556); RR 0.25 (95%CI: 0.13-0.46) mech. ventilation: 0.09% (10/11556) vs. 0.2% (23/11556); RR 0.44 (95%CI: 0.21-0.91)  5-17yo ED visits: 10.09% (1889/18715) vs. 13.07% (2446/18715); RR 0.77 (95%CI: 0.73-0.82) hospitalization: 0.77% (144/18715) vs. 1.37% (144/18715);  RR 0.56 (95%CI: 0.46-0.69) ICU admissions: 0.23% (43/18715) vs. 0.38% (71/18715); RR 0.61 (95%CI: 0.42-0.88) mech. ventilation: 0.05% (10/18715) vs. 0.06 (11/18715); RR 0.91 (95%CI: 0.39-2.14) | **sequencing:** n/r **vaccination status:** n/r |
| Marks  et al. [43] | cohort study | USA | hospital inpatients | infected:  2 100  omicron infection:  266  delta infection:  1 834 | n/r | ≤4 yo, 5-11 yo, 12-17yo | median:  7 yo  (IQR 1-14) | Delta, Omicron | **Severity (omicron vs. delta)** ICU admission (≤17 yo): 20.2% (52/266) vs. 27.8% (510/1834) mech. ventilation (≤17 yo): 2.3% (6/266) vs. 6.3% (112/1834)  hospitalization (≤4yo): 54.2% (142/266) vs. 42.5% (778/1834); RR 5.4 (95%CI: 4.0-7.2)  hospitalization (5-11 yo): 16.9% (43/266) vs. 22.5% (417/1834); RR 2.3 (95%CI: 1.5-3.6) hospitalization (12–17 yo): 28.9% (81/266) vs. 34.9% (639/1834); RR 3.5 (95%CI: 2.5-5.0) | **sequencing:** n/r **vaccination status:** n/r |
| Iuliano  et al. [44] | cross-sectional  study | USA | hospital inpatients | infected: 824  omicron infection: 405  delta infection: 272  non-VOC infection: 147 | n/a | ≤17 yo | n/r | Delta, Omicron | **Severity (omicron vs. delta)** ICU admission: 10.4% (42/405) vs. 18.4% (50/272) (ICU admission non-VOC: 17.0% (25/147)) | **sequencing:** n/r **vaccination status:** n/r |
| Shi  et al. [45] | cross-sectional study | USA | hospital inpatients | infected:  1 475  omicron infection: 397  delta infection: 482  pre-Delta infection:  596 | n/a | 5-11 yo | median:  8 yo  (IQR 6-10) | Delta, Omicron | **Severity (omicron vs. delta)** ICU admission: 18.9% vs. 26.1%, p=0.05  mech. ventilation: 4.6% vs. 6.8%, p=0.28 death: 0% vs. 0%  **Severity (omicron vs. pre-delta)** ICU admission pre-Delta: 18.9% vs. 32.6%, p<0.01 mech. ventilation: 4.6% vs. 6.7%, p=0.28 death: 0%. vs. 0.7%   **underlying diseases (omicron vs. delta):** neurologic disorders: 33% vs. 21%, p<0.01 obesity: 33% vs. 21%, p = 0.01 | **sequencing:** n/r **vaccination status:** n/r |
| Marks et al. [46] | cohort study | USA | Hospital inpatients | infected: 2 562  omicron infection:  572  delta infection:  790  pre-delta infection: 1 200 | n/a | ≤4 yo | median: 0.6yo  (IQR 0.1–1.0) | Delta, Omicron | **Severity  (omicron vs. delta vs. pre-delta):**  length of hospitalisation (days, median, IQR): 1.5 (0.5-2.5) vs. 2 (1-3.5) vs. 1.5 (1-3.5),  p (omicron vs. pre-delta)= 0.001, p (omicron vs. delta)=0.002 ICU admission: 21.0 % vs. 26.7% vs. 24%,  p (omicron vs. pre-delta)= 0.19,  p (omicron vs. delta)=0.02 BiPAP/CPAP^[37]^: 5.1% vs. 9.1% vs. 5.9%,  p (omicron vs. pre-delta)= 0.53,  p (omicron vs. delta)=0.008 high flow nasal canula: 13.4% vs. 20.4% vs. 8.3%, p (omicron vs. pre-delta)= 0.002,  p (omicron vs. delta)=0.002  No difference in invasive mechanical ventilation and in hospital deaths | **vaccination status:** none vaccinated **sequencing:** n/r |

| **Supplementary table 10: Review question 1 - Child/adolescent symptoms/severity of disease when infected with the gamma variant (n=2)** | | | | | | | | | | |
| --- | --- | --- | --- | --- | --- | --- | --- | --- | --- | --- |
| **Author** | **Study design** | **Country** | **Study setting** | **Number of study participants <20 yo^[38]^** | **Number of adult study participants** | **Age groups** | **Mean/Median age** | **VOC^[39]^** | **Symptoms/Severity** | **Comments** |
| Oliveira  et al.  [47] | cohort study | Brazil | hospital inpatients | infected: 21 591  gamma infection: 10 017  non-VOC infection: 11 574 | n/r | <20 yo | mean (gamma): 4.0 yo  (IQR^[40]^ 0.6 - 14.7)  mean  (non-VOC): 5.2 yo  (IQR 1.0 - 14.4) | Gamma | **Severity (gamma vs. non-VOC)** hypoxemia: 52.5% vs. 41.1%, p^[41]^<0.0001 ICU^[42]^ admissions: 28.3% (2498/8830) vs. 24.9% (2745/11037), p<0.0001 invasive vent. support: 10.1% (974/9659) vs. 10.8% (1156/10733) p<0.0001 | **sequencing:** n/r |
| Edward  et al.  [48] | cohort study | USA^[43]^ | hospital outpatients | infected: 499  alpha infection:  96  gamma infection:  38  delta infection:  119  non-VOC infection:  243 | n/a | ≤ 18 yo | median (alpha):  9 yo  (IQR 2-14)  median (gamma):  7 yo  (IQR 1-14)  median (delta):  4 yo  (IQR 1-11)  median  (non-VOC):  7 yo  (IQR 1-12) | Alpha, Delta, Gamma | **Symptoms  (alpha vs. gamma vs. delta vs. non-VOC)** symptomatic: 91.6% (88/96) vs. 89.4% (34/38) vs. 89.9% (107/119) vs. 91.7% (223/243), p=0.92  **Severity - Hospitalization** non-VOC: 3.2% (8/243) reference  alpha: 7.2% (7/96): OR^[44]^ 1.9 (95%CI^[45]^: 0.59-6.3) gamma: 15.7% (6/38):  OR 5.9 (95%: 1.6-21.5), p=0.007 delta: 6.7% (8/119): OR 2.2 (95%CI: 0.69-7.7)  **ICU admission** non-VOC: 1.2% (3/243): reference alpha: 1.0% (1/96):  OR 0.47 (95%CI: 0.02-4.0) gamma: 5.2% (2/38):  OR 2.6 (95%CI: 0.31-17.5) delta: 2.5% (3/119):  OR 1.4 (95%CI: 0.24-9.0)  **Respiratory support** non-VOC: 1.2% (3/243): reference alpha: 0%: OR 0 gamma: 10.5% (4/38):  OR 8.3 (95%CI: 1.5-56.3), p=0.02 delta: 2.5% (3/119)  OR 2.2 (95%CI: 0.34-16.9) | **sequencing:** all sequenced |

| **Supplementary table 11: Review question 3 - Child/Adolescent risk of becoming infected with a SARS-CoV-2 variant of concern (n=15)** | | | | | | | | | | |  |
| --- | --- | --- | --- | --- | --- | --- | --- | --- | --- | --- | --- |
| **Author** | **Study design** | **Country** | **Study setting** | **Number of contacts <20 yo^[46]^** | **Number of adult contacts** | **Age groups** | **Mean/ Median age** | **VOC^[47]^** | **Risk of infection compared to  non-VOCs/other VOCs** | **Risk of infection compared to adults** | **Comments** |
| Schenk  et al. [50] | cohort study | Germany | schools kindergartens crèches | non-VOC: 577   alpha:  756 | non-VOC: 334 alpha: 226 | ≤7 yo, adults | median:  4yo | Alpha | **Risk of infection  (alpha vs. non-VOC)** 0% (0/756) vs.  1.2% (7/577) | **Risk of infection  (children vs. adults)** 0% (0/756) vs.  0% (0/226) | **vaccination status:** n/r **sequencing:** n/r |
| Neuberger et al. [51] | cross-sectional study | Germany | schools kindergartens crèches | n/r 8500 early childhood education and care centres | n/r | ≤2 yo, 4-6 yo, ≥7 yo,  adults | n/r | Alpha | n/a | **Risk of infection  (children vs. adults)**  higher absolute case number,  but overall rate of infection still higher in adults | **vaccination status:** n/r **sequencing:** n/r |
| Nakel et al. [18] | cohort study | Germany | schools kindergartens crèches | 113 | 31 | ≤6 yo | n/r | Alpha | **Risk of infection  (alpha vs. non-VOC)** 15.9% (18/113) vs. 64.7% (22/34) | **Risk of infection** **(children vs. adults)** 15.9% (18/113) vs. 35.5% (11/31) | **vaccination status:** n/r **Sequencing:** few cases sequenced, presence of variant assumed for the rest based on date of sampling and national prevalence |
| Roberts et al. [14] | case-control study | United Kingdom | schools kindergartens crèches | 83 | 34 | ≤4 yo, adults | n/r | Alpha | n/a | **Risk of infection AR**^[48]^ **(children vs. adults)** 25.3% (21/83) vs. 70.6% (24/34)   **AR (children):** - 0-1yo: 44% - 1-2 yo: 31% - 2-3yo: 31% - 3-4 yo: 4% | **vaccination status:** n/r **sequencing:** 1 case sequenced, presence of variant assumed for the rest based on date of sampling and national prevalence |
| Loenenbach  et al. [52] | cohort study | Germany | schools kindergartens crèches | 133 | 60 | ≤6 yo, adults | n/r | Alpha | n/a | **Risk of infection SAR**^[49]^ **overall (children vs. adults)** 21.1% (28/133) vs. 31.7% (19/60)  **SAR per  daycare centre  (children vs. adults)** 1. 31% (15/49) vs. 53% (10/19) 2. 27% (7/26) vs. 28% (5/18) 3. 17% (6/36) vs. 17% (4/23) | **vaccination status:** n/r **sequencing:** n/r |
| Lorthe  et al. [53] | cohort study | Switzerland | schools kindergartens crèches | 70 | 9 | 4-6 yo, adults | n/r | Alpha | n/a | **Risk of infection AR (children vs. adults)** 27.4% (22/70) vs. 22.2% (2/9)  **AR (children by classes)**  1. 62.9% (13/21) 2. 10.5% (2/19) 3. 16.7% (3/18) 4. 13.3% (2/15) | **vaccination status:** n/r **sequencing:** all cases |
| Abu-Raddad et al. [54] | cohort study | Qatar | population | 55 638  <10 yo:  28 791  10-19 yo:  26 847 | 30-39yo:  81 760 | <10 yo, 10-19 yo, adults | n/r | Alpha | n/a | **Risk of infection  (children <10 yo vs. adults)** 1.7% (502/28791) vs. 3.6% (2966/81760) OR^[50]^ 0.35  (95%CI^[51]^: 0.32–0.39)  **Risk of infection  (adolescents  10-19 yo vs. adults)** 3.6% (965/26847) vs. 3.6% (2966/81760) | **vaccination status:** n/r **sequencing:** 85% of all cases sequenced |
| Somekh  et al. [19] | cohort study | Israel | hospital inpatients hospital outpatients | 72 426  Alpha:  50 811  non-VOC: 21 615 | n/r | <10 yo | n/r | Alpha | **Risk of infection (alpha vs. non-VOC)** slope of weekly adjusted incidence: 84.4  (95%CI: 71.1- 97.7) vs 39.1  (95%CI:23.9-54.3) | n/a | **sequencing:** n/r |
| Somekh  et al. [16] | cross-sectional study | Israel | households | Alpha:  77  non-VOC: 58 | n/r | ≤5 yo, 6-17 yo | n/r | Alpha | **Risk of infection  (alpha vs. non-VOC)** ≤5 yo: 72% (18/25) vs. 11.1% (2/18) RR^[52]^ 7.8  (95%CI: 2.1–29.0)  6-17 yo: 75% (39/52) vs. 32.5% (13/40)  RR 2.5  (95%CI: 1.6–4.0) | n/a | **sequencing:** n/r |
| Waltenburg  et al. [17] | cohort study | USA^[53]^ | households | Alpha : 61  non-VOC : 39 | n/r | ≤11 yo, 12-17 yo | Median : 12 yo  (IQR^[54]^ 7-15) | Alpha | **Risk of infection  (alpha vs. non-VOC)** 64% (39/61)  (95%CI: 51-76) vs. 56% (22/39)  (95%CI: 40-72); OR 1.08  (95% CI: 0.40- 2.98) **(≤11 yo vs. 12-17yo)** 78% (21/27)  (95%CI: 58-91) vs. 53% (18/34)  (95%CI: 35-70); OR 1.37  (95%CI:0.61-3.04) | n/a | **vaccination status:** 97% unvaccinated (147/151), 3% partially vaccinated (4/151) **sequencing:** majority of primary cases sequenced |
| Clifford  et al. [55] | cohort study | United Kingdom | households | 67 | 406 | <18 yo, adults | n/r | Delta | n/a | **Risk of infection  (children vs. adults)** RR 0.84 (95%CI: 0.66-0.99)  (risk of being infected by non-elderly adults) | **vaccination status:** n/r **sequencing:** 63.1% of cases sequenced, rest was assumed based on date of sampling and national prevalence |
| Ng  et al.  [56] | cohort study | Singapore | households | 1 136 | 7223 | ≤11 yo, 12-17 yo, adults | median : 36 yo  (IQR 26-51) | Delta | n/a | **Risk of infection SAR (≤11 yo vs.  18-29 yo)** 22.9% (173/757) vs. 12.0% (212/1765) ; aOR^[55]^ 1.43 (95%CI: 1.07-1.93), p=0.017  **SAR (12-17 yo vs. 18-29 yo)** 12.9% (49/379) vs. 12.0% (212/1765) ; aOR 0.97 (95%CI: 0.66-1.42), p^[56]^=0.87 | **vaccination status:**  >12 yo partially vaccinated (results adjusted for vaccination status) **sequencing:** partially sequenced |
| Dougherty  et al. [57] | cohort study | USA | camps,  sport clubs | 122 | 11 | <12 yo,  12-19 yo,  adults | median:  14 yo   (IQR 5-58) | Delta | n/a | **Risk of infection   AR (children vs. adults)**  18.9% (23/122) vs. 27.3% (3/11) | **vaccination status:**  majority unvaccinated/partially vaccinated  **sequencing:** 21 sequenced, rest was assumed based on date of sampling and national prevalence |
| Singana-yagam et al. [58] | cohort study | United Kingdom | community | 30 | 198 | 5-18 yo, adults | median:  41 yo (IQR 28-49) | Delta | n/a | **Risk of infection SAR (children vs. adults)** 40% (12/30) vs. 20.1% (41/198) | **vaccination status:**  70% were fully vaccinated, 23.7% were partially vaccinated, 23% were not vaccinated **sequencing:** partially sequenced, rest was assumed based on date of sampling and national prevalence |

| Lorthe  et al.  [59] | Cohort study | Switzerland | schools kindergartens crèches | 59 | 10 | 3-7 yo, adults | n/r | Omicron |  | **Risk of infection AR (children vs. adults)** 44.1% (26/59) vs. 50% (5/10) **AR (children by classes)** 1. 33.3% (4/12) 2. 15.4% (2/13) 3. 56.3% (9/16) 4. 61.1% (11/18) | **vaccination status:** n/r **sequencing:** none |
| --- | --- | --- | --- | --- | --- | --- | --- | --- | --- | --- | --- |

| **Supplementary table 12: Review question 4 - Risk of child/adolescent transmission when infected with a SARS-CoV-2 variant of concern (n=15)** | | | | | | | | | | | | | |
| --- | --- | --- | --- | --- | --- | --- | --- | --- | --- | --- | --- | --- | --- |
| **Author** | **Study design** | **Country** | **Study setting** | **Number of index cases  <20 yo^[57]^** | **Number of adult index cases** | **number of contacts of children/ adolescents** | **number of contacts of adults** | **Age groups** | **Mean/ Median age** | **VOC^[58]^** | **Risk of transmission compared to non-VOCs/other VOCs** | **Risk of transmission compared to adults** | **Comments** |
| Lindstrom et al. [60] | cohort study | Norway | population | 42  alpha:  14  non-VOC: 28 | 373  alpha:  131  non-VOC: 242 | mean (SD^[59]^):  2.79 (1.63) | 40-59yo: 1.72 (1.87)  20-39yo: 1.03 (1.36)  >60yo:  1.25 (1.65) | <20 yo, adults | n/r | Alpha | n/a | **Transmission risk  SAR**^[60]^ **(<20 yo vs.  40-59 yo)** ref. vs. 1.97 (1.08-3.87)  no significant difference to adults  20-39 yo, >60yo | **vaccination status:** n/r **sequencing:** primary cases sequenced, variants in contacts assumed based on date of sampling and national prevalence |
| Lyngse  et al. [61] | cohort study | Denmark | households | 145  <10 yo: 54  10-19 yo: 91 | 663 | 383  <10yo:  141  10-19yo: 242 | 1 336 | ≤10 yo, 10-20 yo, adults | n/r | Alpha | n/a | **Transmission risk AR**^[61]^ **(≤20 yo vs. adults)** 32.4% (124/383) vs. 39.8% (532/1336)  **AR  (≤10 yo vs.  10-19 yo vs. adults)** 45% (64/141) vs. 25% (60/242) vs. 39.8% (532/1336) | **vaccination status:** n/r **sequencing:** 75% sequenced, rest was assumed based on date of sampling and national prevalence |
| Chudasama  et al.  [62] | cohort study | United Kingdom | households | 6 011  <10 yo:  1 289  10-19 yo: 4 722 | 57 382 | n/r | n/r | <10 yo, 10-19 yo, adults | n/r | Alpha | n/a | **Transmission risk OR^[62]^  (10-19 yo vs. <10 yo)**  OR 0.83 (95%CI^[63]^:  0.69-0.99) (<10 yo: reference)  no difference in transmission risk compared to index cases aged 30-70 yo | **vaccination status:** n/r **sequencing:** all index cases sequenced, no information on contacts |
| Julin  et al.  [63] | cohort study | Norway | households | 3  alpha:  2  non-VOC: 1 | 55  alpha:  16  non-VOC: 39 | 55  alpha:  16  non-VOC:  39 | 68  alpha:  20  non-VOC: 48 | <20 yo, adults | median:  31 yo  (IQR^[64]^ 2-73) | Alpha | n/a | **Transmission risk (irrespective of VOC) Viral loads (mean, <20 yo vs. adults)** 2.09 log10 copies/ul RNA^[65]^ vs. 2.98 log10 copies/ul RNA   **Length of testing positive (mean days, <20 yo vs. adults)** 11.3 (95%CI: 7.6–15.1) vs 16.4 (95%CI: 13.5–19.3),  p^[66]^ = 0.03 | **vaccination status:**  none vaccinated **sequencing:** all sequenced |

| Loenenbach et al. [52] | cohort study | Germany | schools kindergartens crèches households | 22 | 16 | 59 | 33 | ≤6 yo, adults | n/r | Alpha | n/a | **Transmission risk SAR^[67]^  (≤6 yo vs. adults)** 39% (95%CI: 28-52) vs. 33% (95%CI: 20-50) RR^[68]^ 1.17 (95%CI: 0.66-2.09) | **vaccination status:** n/r **sequencing:** n/r |
| --- | --- | --- | --- | --- | --- | --- | --- | --- | --- | --- | --- | --- | --- |
| Lorthe et al. [53] | cohort study | Switzerland | schools kindergartens crèches households | n/r | n/r | 24 | 2 | ≤6 yo, adults | n/r | Alpha | n/a | **Transmission risk SAR  (≤6 yo vs. adults)** 12.5% (3/24) vs. 50% (1/2) | **vaccination status:** n/r **sequencing:** all cases |
| Loss  et al. [64] | case-control study | Germany | schools kindergartens crèches households | 15  alpha: 9  non-VOC: 6 | 9  alpha: 4  non-VOC: 5 | 172  alpha:  114  non-VOC: 58 | 38 | ≤6 yo, adults | n/r | Alpha | **Transmission risk AR^[69]^  (alpha vs.  non-VOC)** 16.7% (19/114) vs. 8.6% (5/58) | **Transmission risk AR  (≤6 yo vs. adults)** 16.7% (19/114) vs. 10.5% (4/38) | **vaccination status:** n/r **sequencing:** majority sequenced, rest was assumed based on date of sampling and national prevalence |
| Buchan  et al. [65] | cohort study | Canada | households | 715  <10 yo alpha: 55 non-VOC:  195  10-19 yo alpha: 114 non-VOC: 351 | 4 902  alpha:  1 149 non-VOC:  3 753 | n/r | n/r | <10 yo, 10-19 yo, adults | n/r | Alpha | **Transmission risk SAR  (alpha vs.  non-VOC)** aRR^[70]^ (<10y o): 1.47 (95%CI: 0.81-2.65)  aRR  (10-19 yo): 1.30 (95%CI: 0.80-2.10) | n/a | **sequencing:** all cases either sequenced or screened positive for N501Y (marker for alpha variant) by PCR^[71]^ |
| Waltenburg et al.  [17] | cohort study | USA^[72]^ | households | 36  alpha:  21  non-VOC: 10 | n/a | 111  alpha : 61  non-VOC :  39 | n/a | <5 yo 5-11 yo 12-17 yo | median: 12 yo  (IQR 7-15) | Alpha | **Transmission risk  SAR  (alpha vs.  non-VOC)** 55% (24/44) (95%CI: 39-70) vs. 46% (12/26) (95%CI: 27-67); OR 1.52 (95%CI:  0.51-4.53) **SAR – alpha (≤11 yo vs.  12-17 yo)** 65% (11/17) (95%CI: 38-68) vs. 48% (13/27) (95%CI: 29-68); OR 1.63 (95%CI:  0.4-6.7) | n/a | **vaccination status:** 97% unvaccinated (147/151), 3% partially vaccinated (4/151) (2 primary cases and 2 secondary cases)  **sequencing:** majority of primary cases sequenced |
| Ratman  et al.  [66] | modeling study | USA | population | n/a | n/a | n/a | n/a | <10 yo, 10-19 yo | n/r | Alpha | **Transmission risk R**^[73]^ **(alpha vs. non-VOC)** <10yo: <1 vs. <<1 10-19 yo: ~1 vs. <<1 | n/a | n/a |
| Somekh et al. [19] | cohort study | Israel | hospital inpatients hospital outpatients | 72 426  alpha:  50 811  non-VOC: 21 615  (overall numbers and thus potential index cases) | n/r | alpha:  313 871  non-VOC: 156 521 | n/r | <10 yo | n/r | Alpha | **Transmission risk AR  (alpha vs.  non-VOC)** 15.7% (49257/313871) vs. 7.5% (11770/156521)  RR: 2.24  (95%CI, 2.20-2.29), p < 0.001 | n/a | **sequencing:** n/r |
| Allen et al.  [67] | case-control  study | United Kingdom | households | 6 130  alpha: n/r  delta: n/r | 17 928 | n/r | n/r | <10 yo, 10-19 yo, adults | n/r | Alpha, Delta | n/a | **Trasmission risk aOR^[74]^ (delta vs. alpha/ref. adults)** <10 yo: 0.98 (95%CI:  0.85-1.14)  10-19 yo: 0.71 (95%CI:  0.64-0.79)  (reference value 1.00 for 30-39 yo) | **vaccination status:** reported (but not in subanalysis comparing transmission risk in age groups)  **sequencing:** all index cases sequenced, nationwide 50% of all cases were sequenced at time of study |
| Trobajo-Sanmartin et al. [68] | cohort study | Spain | population | n/r | n/a | All contacts of <12 yo : 288  Alpha : 107  Delta : 181 | n/a | <12yo, 12-17 yo, adults | n/r | Alpha, Delta | **Transmission risk  SAR (alpha vs. delta)** <12 yo : 33% (35/107) vs. 23% (41/181); aRR 1 vs. 0.58 (95%CI: 0.28-1.23), p=0.155  higher risk of transmission for 12-39yo |  | **vaccination status:** none vaccinated **sequencing:** none |
| Ng  et al. [56] | cohort study | Singapore | households | n/r | n/r | 472 | 18-29 yo :  1 467 | ≤11 yo, 12-17 yo, adults | median (contacts): 36 yo  (IQR 26-51) | Delta | n/a | **Transmission risk** **SAR  (≤11 yo vs.  18-29 yo)** 25.2% (88/349) vs. 9.4% (138/1457); aOR: 2.37 (95%CI:1.57-3.60), p<0.0001  **SAR  (12-17 yo vs. 18-29 yo)** 17.9% (22/123) vs. 9.4% (138/1457) ;  aOR 1.81 (95%CI: 0.91-3.6), p=0.089 | **vaccination status:**  >12 yo partially vaccinated (results adjusted for vaccination status) **sequencing:** partially sequenced |
| Lorthe  et al. [59] | cohort study | Switzerland | schools kindergartens crèches, households | n/r | n/a | 52 | n/a | 3-7 yo, adults | n/r | Omicron | **Transmission risk  SAR** 48% (25/52) | n/a | **vaccination status:** n/r  **sequencing:** none |

1. yo: years old
2. VOC: Variants Of Concern
3. RR: Risk Ratio
4. 95%CI: 95% Confidence Interval
5. USA: United States of America
6. IQR: Interquartile Range
7. OR: Odd Ratio
8. p: p-value
9. ICU: Intensive Care Unit
10. ^6^PIMS-TS/MIS-C: Paediatric Inflammatory Multisystem Syndrome/Multisystem Inflammatory Syndrome in Children
11. yo: years old
12. VOC: Variants Of Concern
13. SD: Standrad Deviation
14. MIS-C: Multisystem Inflammatory Syndrome in Children
15. ICU: Intensive Care Unit
16. ECMO: Extracorporeal Membrane Oxygenation
17. RR: Risk Ratio
18. p: p-value
19. OR: Odd Ratio
20. IQR: Interquartile Range
21. USA: United States of America
22. aOR: Adjusted Odd Ratio
23. RR: Risk Ratio
24. 95%CI: 95% Confidence Interval
25. yo: years old
26. VOC: Variants Of Concern
27. USA: United States of America
28. ED: Emergency Department
29. IQR: Interquartile Range
30. p: p-value
31. aOR: Adjusted Odd Ratio
32. COVID: Coronavirus Disease
33. OR: Odd Ratio
34. 95%CI: 95% Confidence Interval
35. RR: Risk Ratios
36. ICU: Intensive Care Units
37. BiPAP/CPAP: Bilevel Positive Airway Pressure/ Continuous Positive Airway Pressure
38. yo: years old
39. VOC: Variant Of Concern
40. IQR: Interquartile Range
41. p: p-value
42. ICU: Intensive Care Unit
43. USA: United States of America
44. OR: Odds Ratio
45. 95%CI: 95% Confidence Interval
46. yo: years old
47. VOC: Variant Of Concern
48. AR : Attack Rate
49. SAR: Secondary Attack Rate
50. OR: Odd Ratio
51. 95%CI: 95% Confidence Interval
52. RR: Risk Ratio
53. USA: United States of America
54. IQR: Interquartile Range
55. aOR: adjusted Odd Ratio
56. p: p-value
57. yo: years old
58. VOC: Variant Of Concern
59. SD: Standard Deviation
60. SAR: secondary attack rate
61. AR: attack rate
62. OR: Odd Ratio
63. 95%CI: 95% Confidence Interval
64. IQR: Interquartile Range
65. RNA: Ribonucleic acid
66. p: p-value
67. SAR: Secondary Attack Rate
68. RR: Risk Ratio
69. AR: Attack Rate
70. aRR: Adjusted Risk Ratio
71. PCR: Polymerase Chain Reaction
72. USA: United States of America
73. R: reproduction number
74. aOR: Adjusted Odd Ratio
