## Supplementary figure 1 - flowchart 1 for "The role of SARS-CoV-2 variants of concern in children and adolescents with COVID-19: a systematic review"

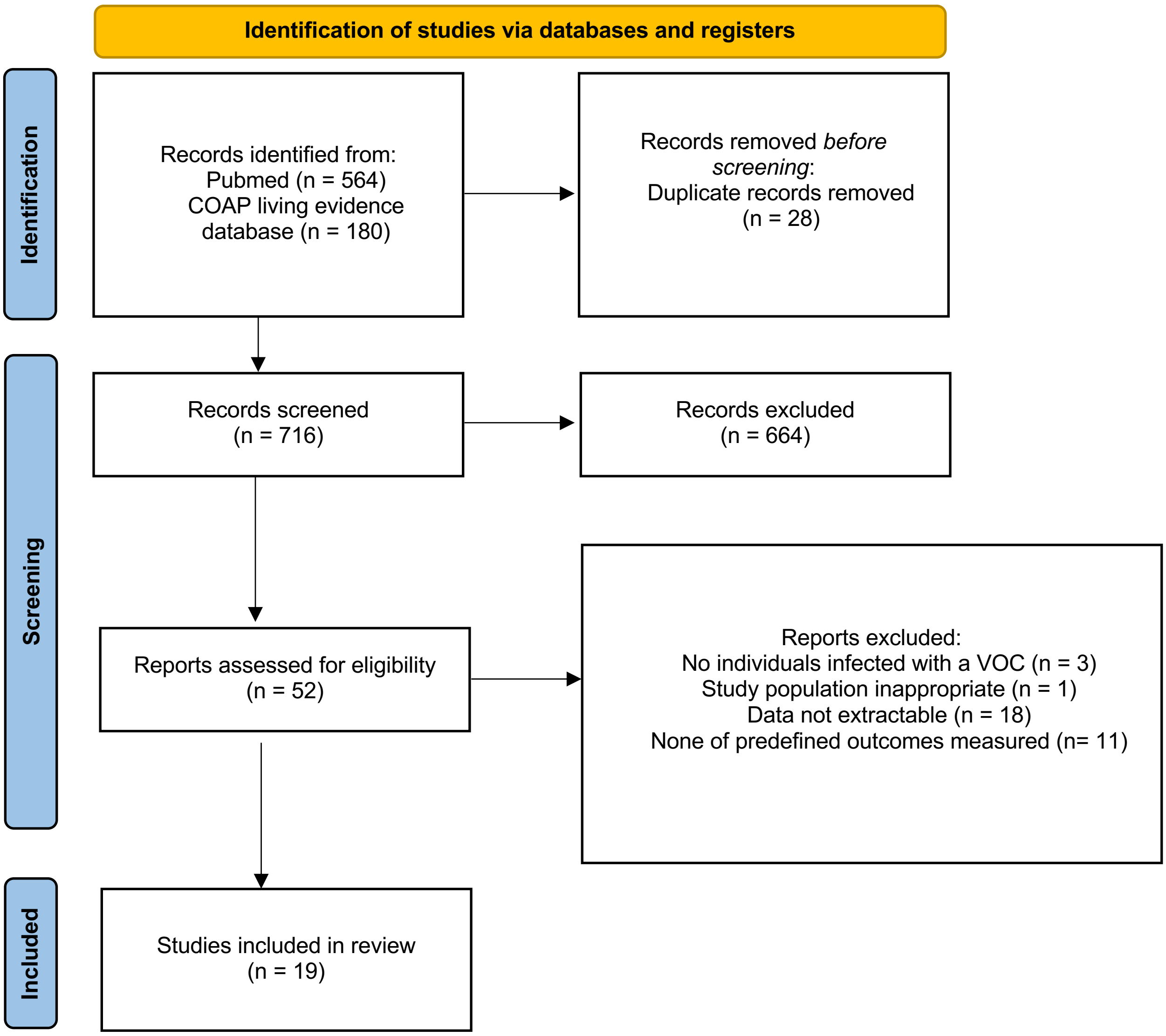


**Supplementary figure 1: Flow chart – first search (until 15.10.2021)**
Of the 744 studies found via database searches 19 were found to be eligible for this systematic review.

*From:* Page MJ, McKenzie JE, Bossuyt PM, Boutron I, Hoffmann TC, Mulrow CD, et al. The PRISMA 2020 statement: an updated guideline for reporting systematic reviews. BMJ 2021;372:n71. doi: 10.1136/bmj.n71
