## Supplementary figure 2 - flowchart 2 for "The role of SARS-CoV-2 variants of concern in children and adolescents with COVID-19: a systematic review"

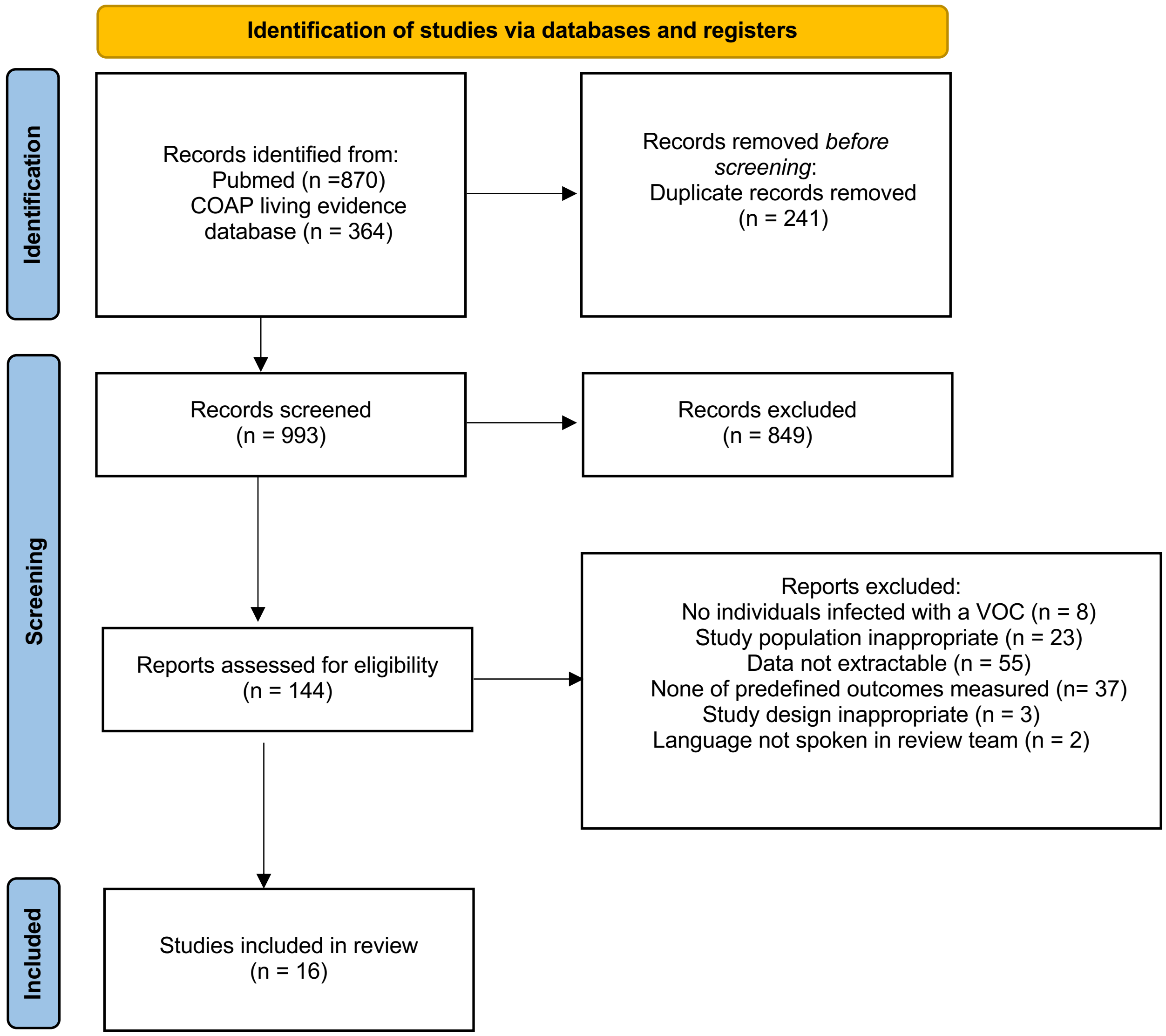


**Supplementary figure 2: Flow chart – second search (15.10.2021-31.01.2022)**
**Of the 1234 studies found via database searches 16 were found to be eligible for this systematic review.**

*From:* Page MJ, McKenzie JE, Bossuyt PM, Boutron I, Hoffmann TC, Mulrow CD, et al. The PRISMA 2020 statement: an updated guideline for reporting systematic reviews. BMJ 2021;372:n71. doi: 10.1136/bmj.n71
