## Supplementary figure 4 - flowchart 4 for "The role of SARS-CoV-2 variants of concern in children and adolescents with COVID-19: a systematic review"

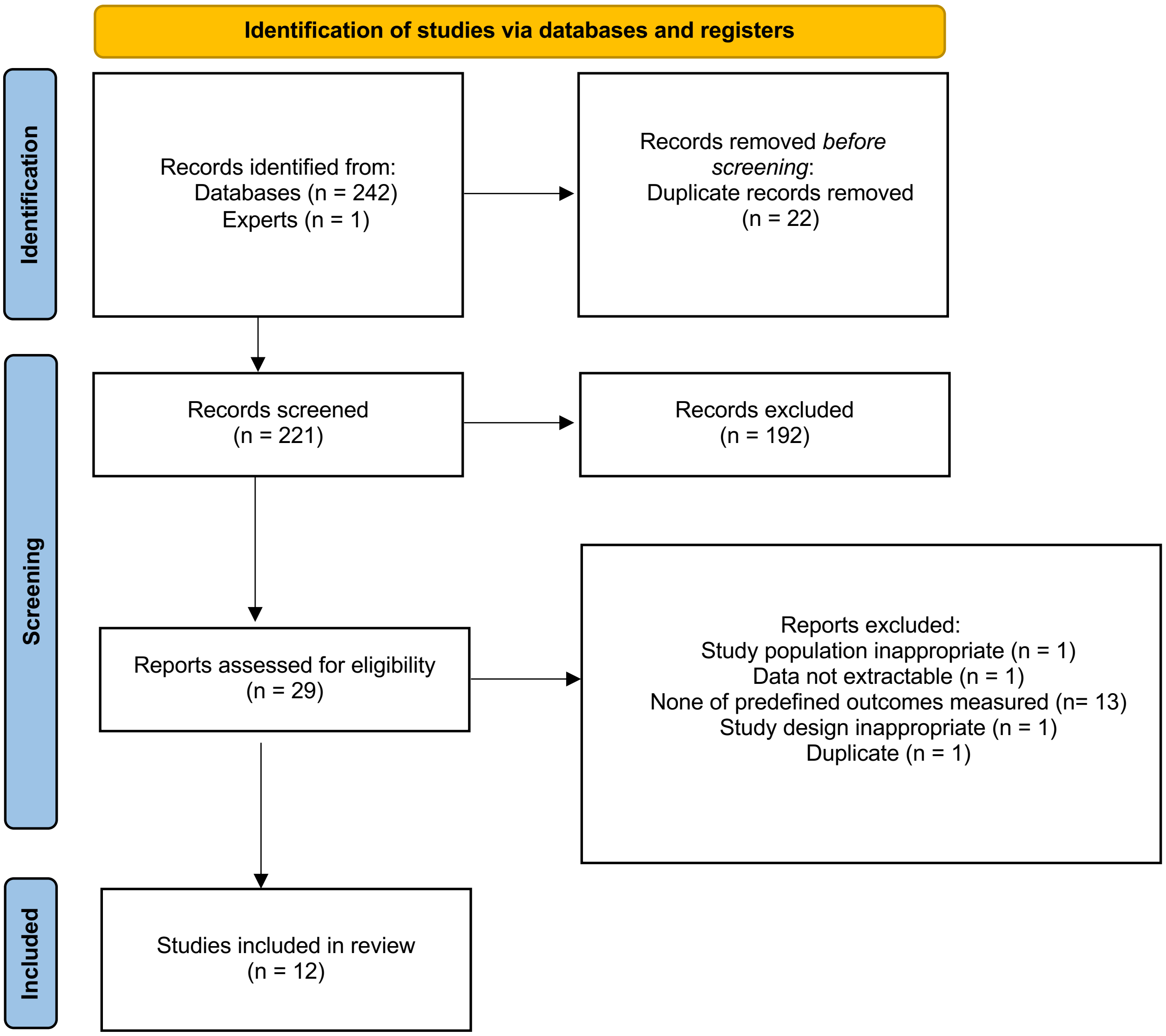


**Supplementary figure 4: Flow chart – fourth search (07.03.2022-09.05.2022)**
**Of the 243 studies found via the database search (incl. 1 study recommended by experts) 12 studies were found to be eligible.**

*From:* Page MJ, McKenzie JE, Bossuyt PM, Boutron I, Hoffmann TC, Mulrow CD, et al. The PRISMA 2020 statement: an updated guideline for reporting systematic reviews. BMJ 2021;372:n71. doi: 10.1136/bmj.n71

For more information, visit: http://www.prisma-statement.org/
