## Supplementary note 1 - search strategy for "The role of SARS-CoV-2 variants of concern in children and adolescents with COVID-19: a systematic review"

**URL to search strategy**

a) COAP database search terms: (child* AND variant*) in title or abstract
b) PubMed search child*"[Title/Abstract] OR "neonat*"[Title/Abstract] OR "adolesc*"[Title/Abstract] OR "infan*"[Title/Abstract] OR "teen*"[Title/Abstract] OR "pediatr*"[Title/Abstract] OR "paediatr*"[Title/Abstract] OR ("educational status"[MeSH Terms] OR ("educational"[Title/Abstract] AND "status"[Title/Abstract]) OR "educational status"[Title/Abstract] OR "schooling"[Title/Abstract] OR "education"[MeSH Terms] OR "education"[Title/Abstract] OR "school s"[Title/Abstract] OR "schooled"[Title/Abstract] OR "schools"[MeSH Terms] OR "schools"[Title/Abstract] OR "school"[Title/Abstract]) OR ("nurseries, infant"[MeSH Terms] OR ("nurseries"[Title/Abstract] AND "infant"[Title/Abstract]) OR "infant nurseries"[Title/Abstract] OR "nurseries"[Title/Abstract] OR "nursery"[Title/Abstract]) OR ("infant, newborn"[MeSH Terms] OR ("infant"[Title/Abstract] AND "newborn"[Title/Abstract]) OR "newborn infant"[Title/Abstract] OR "newborn"[Title/Abstract] OR "newborns"[Title/Abstract] OR "newborn s"[Title/Abstract]) OR "toddler*"[Title/Abstract] OR ("minority groups"[MeSH Terms] OR ("minority"[Title/Abstract] AND "groups"[Title/Abstract]) OR "minority groups"[Title/Abstract] OR "minorities"[Title/Abstract] OR "minority"[Title/Abstract] OR "minority s"[Title/Abstract] OR "minors"[MeSH Terms] OR "minors"[Title/Abstract] OR "minor"[Title/Abstract])
   AND
   “COVID-19”[tiab] OR 2019-novel-cov*[tiab] OR coronavirus[tiab] OR coronavirus-disease-19*[tiab] OR corona-virus-disease-19*[tiab] OR covid-19*[tiab] OR covid19*[tiab] OR new-coronavirus[tiab] OR new-corona-virus[tiab] OR novel-coronavirus[tiab] OR novel-corona-virus[tiab] OR sars-2*[tiab] OR sars2*[tiab] OR sars-cov-19*[tiab] OR sarscov19*[tiab] OR sars-cov-2*[tiab] OR sars-cov2*[tiab] OR sarscov2*[tiab] OR sarscov-2*[tiab]
   AND
   "VOC"[All Fields] OR "variant of concern"[All Fields] OR "alpha variant"[All Fields] OR "beta variant"[All Fields] OR "gamma variant"[All Fields] OR "delta variant"[All Fields] OR “omicron variant”[All Fields] OR "variant of interest"[All Fields]
